# Detecting Potential Mediators of the Causal Effect of Education Level on The Risk of Stroke: A Two-Step, Two-Sample Multivariable Mendelian Randomization Study

**DOI:** 10.1101/2024.10.23.24316024

**Authors:** C Ken, Z Ying, W Zheng, Y Ying

## Abstract

**Background:** The effect of education level on the risk of stroke is not fully understood. The present study aimed to reveal the genetic and modifiable mediators for the effect of education level on the risk of stroke.

**Methods:** Summary-level genetic data were obtained from GWAS consortia. Two-sample Mendelian Randomization (MR) analysis was performed to uncover the causal effect of education level on the risk of stroke, and regression-based multivariable MR (MVMR) analyses were used to explore potential mediators.

**Results:** Genome-wide MR analyses showed that genetically determined higher education level was associated with reduced risk of stroke (Odds ratio (OR) per standard deviation (SD) increase: 0.74[95% confidence interval (CI): 0.58 to 0.94]; p = 0.013). We found instant coffee intake, never smoking status, body mass index (BMI), waist circumference (WC), waist-hip-ratio (WHR), Type-2 diabetes mellitus (T2DM), Diastolic pressure (DBP), Systolic pressure (SBP), Alanine aminotransferase (ALT), Platelet (PLT) and CTRP-1 (complement c1q tumor necrosis factor-related protein 1) had significant mediation roles in the effect of education level on the risk of stroke. The proportion of total effect mediated by these modifiable factors was 79%.

**Conclusion:** We found many modifiable mediators had essential mediation roles in the effect of education level on the risk of stroke. Intervention on these mediators might have protective effect on the risk of stroke, which highlighted novel therapeutic targets.

## Introduction

Stroke remains a serious public health concern globally. According to recent epidemiological investigations, nearly 10 million new cases of stroke occurred globally each year, including 1.5 million in Europe (1), and the prevalence of stroke was 2.5% in the US. Moreover, the incidence of stroke is even higher in Asia (2). Stroke also inflicted an enormous economic and social burden (2, 3). Improvement of recognition and control of risk factors offered promising therapeutic targets, but there is still a long way to go for more effective preventive and therapeutic strategies.

Recent studies have showed that apart from traditional cardiovascular risk factors, education level was another independent risk factor, and low education level has been associated with increased risk of stroke (4, 5). However, results of available observational studies could be influenced by confounders and other unmeasured or unknown factors, thus were less effective for proving the causal relationship of education level with stroke. Besides, it is not feasible to conduct randomized controlled trials (RCTs) to explore the causal relationship between education level and stroke. Moreover, the potential pathways from education level to stroke remained unestablished.

Mendelian Randomization (MR) is an established epidemiological method, which use genetic variants as instrumental variables to explore putative causal effects of exposures on outcomes of interest (6). Since genotype is allocated randomly at conception, MR studies are less liable to confounding factors or measurement errors (7). The two-sample MR method indicates that genetic variants for exposures and outcomes are obtained from different datasets, which allows it to be more robust in statistic power, but overlap between these two datasets may cause alternative source of bias (8).

Previous MR studies indicated that a higher education level seemed to be protective against ischemic stroke (9), (10). Yet the underlying mechanism of the protective effect remained not well understood, and whether intervention on some modifiable factors could mitigate the risk of stroke was also unclear. Therefore, we conducted a two-step, two-sample multivariable MR (MVMR) study to further explore the possible mediators of the effect of education level on stroke. We quantified how much of the effect of education level on stroke were mediated through lifestyle modification, cardiometabolic factors and other biomarkers etc. by analyzing genome-wide association study (GWAS) summary statistics from international genetic consortia.

## Methods

The present study was exempted from ethical approval because it used summary-level data. Our Study was reported according to the STROBE-MR guidelines (Strengthening the Reporting of Observational Studies in Epidemiology-Mendelian Randomization).

### Overall Study Design

The first step of our two-step MR study is to uncover the causal effect of education level on the risk of stroke. The second step is to explore possible modifiable mediators of the effect of education level on the risk of stroke using MVMR method.

### Data Sources

#### Genetic Instrumental Variables for Education

For most people, higher education levels require longer periods of study on campus which can lead to older age completed full-time education. Thus, we obtained over 9 million genetic variants for age completed full-time education from an online database of UK Biobank (UKB) which consists of 307,897 European ancestry through R software (R Consortium, Boston, MA) TwoSampleMR package (http://gwas-api.mrcieu.ac.uk/).

#### Genetic Instrumental Variables for Potential Mediators

After thorough screening, we selected 19 potential mediators including lifestyle behaviors (instant coffee consumption, insomnia, physical activity, never smoking status, alcohol intake), cardiometabolic factors (body mass index (BMI), WC (waist circumference), waist-hip-ratio (WHR), type-2 diabetes mellitus (T2DM), systolic blood pressure (SBP), diastolic blood pressure (DBP), high-density lipoprotein (HDL), low-density lipoprotein (LDL), triglyceride (TG)), other biomarkers (alanine aminotransferase (ALT), aspartate aminotransferase (AST), Platelet (PLT), complement c1q tumor necrosis factor-related protein 1 (CTRP-1)) and depression. Genetic variants for instant coffee consumption, insomnia, physical activity, never smoking status, alcohol intake, WC, DBP, level of ALT, AST, level of PLT were extracted from different online databases of UKB provided by Neale lab and Ben Elsworth (http://gwas-api.mrcieu.ac.uk/). Genetic variants for BMI were drawn from a GWAS including 681,275 European ancestry (11). We obtained genetic variants for WHR from a meta-analysis including 212,244 European ancestry (12). We acquired genetic data for T2DM from a meta-analysis of GWAS which consists of over 16 million genetic variants of European ancestry (13). For SBP, genetic data were extracted from a genetic analysis including over one million people drawn from UKB with 458,577 European participants (14) and the International Consortium of Blood Pressure-Genome Wide Association Studies (ICBP) (15, 16), consisting of 299,024 European individuals. Genetic variants for HDL, LDL, and TG were extracted from a GWAS including 188,577 participants which were mainly European ancestry, respectively (17). Genetic variants for depression were obtained from a GWAS with 322,580 European participants (18). We extracted genetic variants for CTRP-1 from a genetic analysis in 3,301 individuals of European descent (19).

#### Genetic Instrumental Variables for Stroke

Genetic summarized statistics for stroke were obtained from the MEGASTROKE Consortium with over 520,000 subjects originating from mixed ancestry (mostly were European) (20). Instrumental variables for stroke included genetic variants for ischemic stroke (IS), intracerebral hemorrhage (ICH), large-artery atherosclerosis (LAS), cardioembolic stroke (CES), and stroke caused by small-vessel disease (SVS).

Details of data sources for age completed full-time education, potential mediators and stroke were shown in Supplementary Table 1. All the genetic variants used as instrumental variables were shown in Supplementary Table 2-21.

### Statistical Analysis

#### Instrumental Variable Selection

We performed thorough screening and chose genetic instruments at the level of genome-wide significance (P < 5×10^-8^). Besides, for genetic instruments in linkage disequilibrium (LD), we kept one instrument with the lowest P-value. F-statistic which is calculated using the equation (β/SE)^2^, is commonly applied in MR studies to evaluate the strength of genetic instruments.

#### Effect of Education on Stroke

The effect of education level on the risk of stroke was estimated using a two-sample MR design. Random effect model of inverse-variance weighted (IVW) method was applied to estimate the causal effect of education level on the risk of stroke. Results were shown using odds ratio (OR) or β coefficient and 95% confidence interval (CI). P<0.05 for the IVW method was considered suggestive for potential causal association.

#### Effects of Education on Mediators

We applied a two-sample MR design to estimate the effect of education on each potential mediator. Results were shown using OR or β coefficient and 95% CI. Random effect model of IVW method was applied as main analysis and P < 0.05 for the IVW method was considered suggestive for potential causality. Genetic variants for HDL, LDL and TG were extracted from the same GWAS database, therefore, a Bonferroni corrected p-value threshold less than 0.017 was applied to correct for multiple testing, and Bonferroni corrected p < 0.017 was considered statistically significant.

#### Effects of Mediators on Stroke

Estimates of the effects of potential mediators on stroke adjusting for education were obtained by regression based multivariable MR (MVMR) (21). Results were shown using OR and 95% CI. For those did not meet p < 0.05 standard were considered not statistically significant, thus were excluded from subsequent analysis.

#### Mediation Effects of Potential Mediators

To obtain the mediation effect of each potential mediator individually, the estimate of the effect of education level on each potential mediator was multiplied with the estimate of the effect of each mediator on stroke, respectively. Then we divided the individual mediation effect by the total effect of education level on stroke. All analyses were performed using R version 4.0.3 (The R Foundation for Statistical Computing) through the TwoSampleMR package and Mendelian Randomization package. To fit different situations such as multiplication and division, SEs were calculated using rules which were derived from the Gaussian equation (Supplementary Methods).

#### Sensitivity Analyses

Several sensitivity analyses were applied in our study, including weighted median, MR-Egger regression, MR-PRESSO to explore potential bias from potential pleiotropy. We also applied single nucleotide polymorphism (SNP) analysis, leave-one-out analysis to investigate the influence of possible outlying SNPs. Full details of these sensitivity analyses were shown in Supplementary Methods.

## Results

After ruling out SNPs which did not meet the standard of genome-wide significance (p < 5 × 10^-8^) and clumping for linkage disequilibrium (LD) (r^2^ < 0.001), we identified 41 SNPs for age completed full-time education. The F-statistics of these selected SNPs were larger than 10, which indicated that results of our study were less likely influenced by weak instruments (22).

### Total Effect of Education on Stroke

Strong evidence supporting the causal effect of education on stroke was identified in our study. Figure 1 showed the total effect of education level on stroke. One-unit higher log odds of age completed full-time education reduced 30% risk of stroke. (OR: 0.74, 95% CI: 0.58-0.94, p <0.05). Details of genetic associations of education on stroke were shown in Supplementary Table 22.

**Figure 1.**
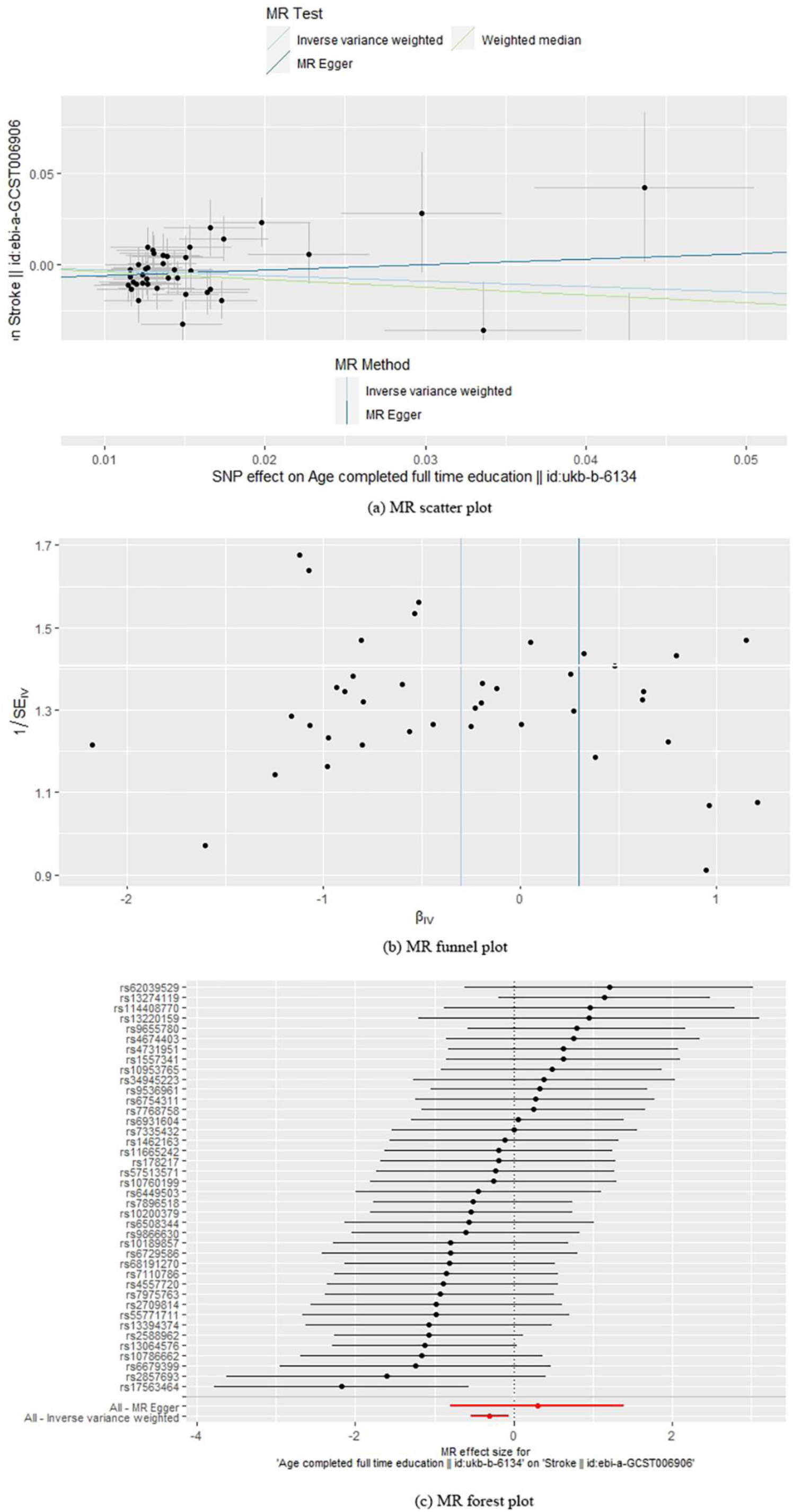
Estimate of effect of age completed full-time education on stroke. (a). MR scatter plot; (b). MR funnel plot; (c). MR forest plot. MR: Mendelian Randomization.

### Effect of Education on Potential Mediators

Figure 2 showed one-unit higher log odds of age completed full-time education was associated with higher probability of never smoking status (OR: 1.13, 95% CI: 1.07-1.20, p < 0.001), higher rate of physical activity (OR: 1.15, 95% CI: 1.10-1.20, p < 0.001), more alcohol intake (OR: 1.39, 95% CI: 1.32-1.46, p < 0.001), less insomnia (OR: 0.86, 95% CI: 0.82-0.91, p < 0.001), less instant coffee intake (OR: 0.86, 95% CI: 0.83-0.89, p < 0.001), lower probability of T2DM (OR: 0.51, 95% CI: 0.37-0.71, p < 0.001), less rate of depression (OR: 0.92, 95% CI: 0.88-0.97, p = 0.003), higher level of HDL (β: 0.24, 95% CI: 0.06 to 0.43, p = 0.011), lower level of BMI (β: -0.31, 95% CI: -0.45 to -0.17, p < 0.001), WC (β: -0.28, 95% CI: -0.37 to - 0.19, p < 0.001), WHR (β: -0.31, 95% CI: -0.47 to -0.15, p < 0.001), DBP (β: -0.31, 95% CI: - 0.43 to -0.19, p < 0.001), SBP (β: -2.95, 95% CI: -4.77 to -1.14, p = 0.001), TG (β: -0.38, 95% CI: -0.57 to -0.19, p < 0.001), ALT (β: -0.15, 95% CI: -0.24 to -0.06, p = 0.002), AST (β: -1.06, 95% CI: -1.97 to -0.15, p = 0.020), PLT (β: -0.18, 95% CI: -0.35 to -0.02, p = 0.030), CTRP-1 ((β: -0.66, 95% CI: -1.31 to -0.01, p = 0.047). We did not detect genetic association between education level and the level of LDL (p>0.05). Genetic association of education on each potential mediator was shown in Supplementary Table 23-41. Effects of Potential Mediators on Stroke Figure 3 showed the estimate of effect of 1-SD or 1-unit higher log odds of each potential mediator on stroke after adjusting for education level. Estimate of log odds of stroke for 1-SD increase in the level of BMI, WC, WHR, DBP, SBP, CTRP-1, ALT and PLT was 1.10 (95% CI: 1.02 to 1.18, p = 0.010), 1.15 (95% CI: 1.03 to 1.29, p = 0.015), 1.20 (95% CI: 1.03 to 1.40, p = 0.030), 1.64 (95% CI: 1.45 to 1.85, p < 0.001), 1.03 (95% CI: 1.03 to 1.04, p < 0.001), 1.03 (95% CI: 1.02 to 1.05, p = 0.010), 1.15 (95% CI: 1.06 to 1.26, p = 0.002), 1.08 (95% CI: 1.04 to 1.14, p < 0.001), respectively. 1-unit higher log odds of instant coffee intake, never smoking status, and T2DM was also causally associated with the risk of stroke, the estimate of log odds of stroke was 0.48 (95% CI: 0.45 to 0.51, p = 0.030), 0.53 (95% CI: 0.34 to 0.84, p = 0.008), 1.09 (95% CI: 1.06 to 1.13, p < 0.001), respectively. For mediators including insomnia, physical activity, alcohol intake, depression and level of HDL, TG, AST, p-values of the estimates of these mediators on stroke were larger than 0.05, which indicated the association between these mediators and stroke did not meet statistical significance, thus, we did not take these mediators into final mediation analyses. Details of the effects of potential mediators on stroke were shown in Supplementary Table 43-61.

**Figure 2.**
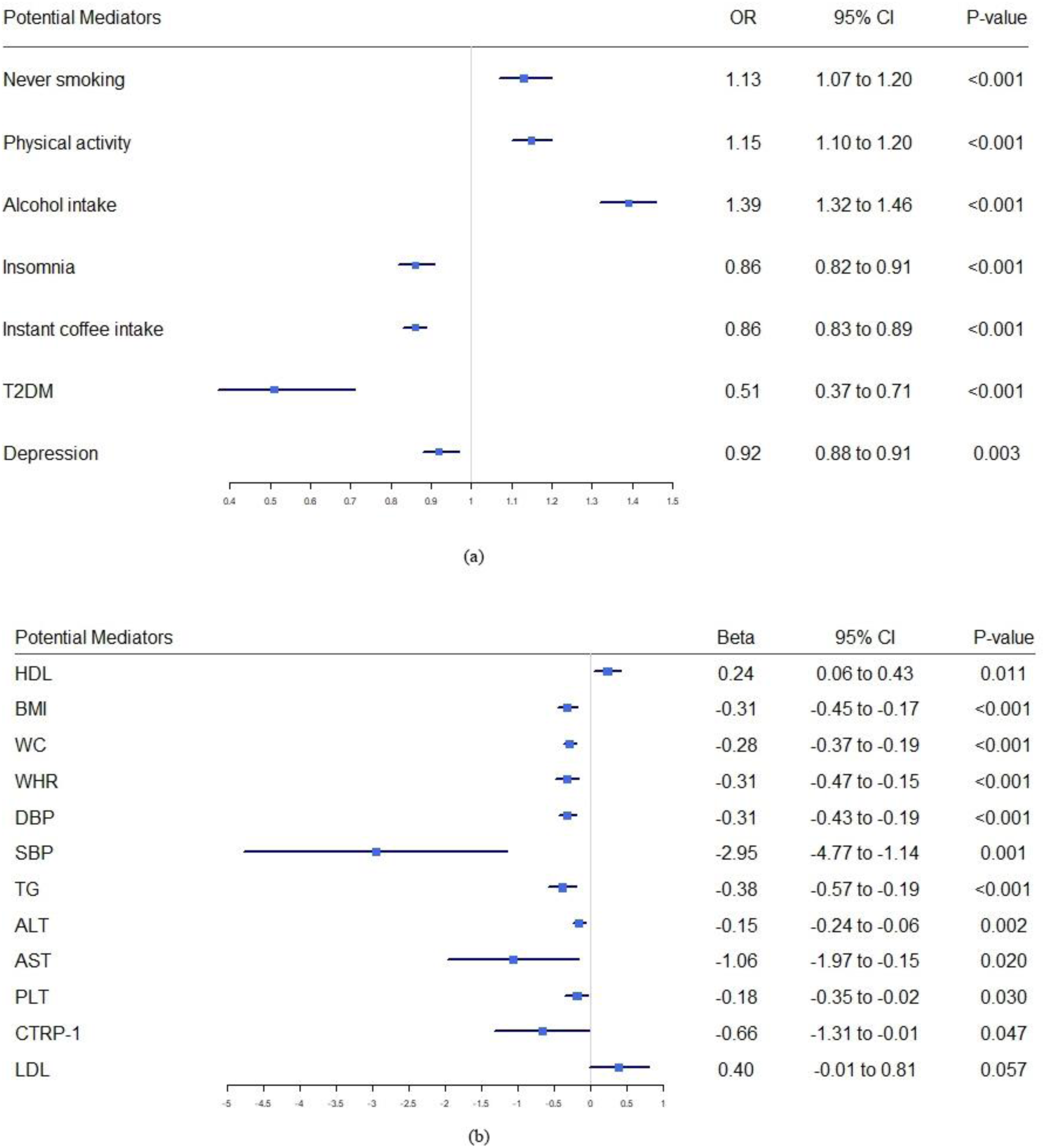
(a). Estimate of effect of one-unit higher log odds of age completed full-time education on never smoking status, physical activity, alcohol intake, insomnia, instant coffee intake, T2DM and depression. (b). Estimate of effect of one-unit higher log odds of age completed full-time education on the level of HDL, BMI, WC, WHR, DBP, SBP, TG, ALT, AST, PLT, CTRP-1 and LDL. OR: Odds ratio; CI: Confidence interval; T2DM: Type-2 diabetes; HDL: High-density lipoprotein; BMI: Body mass index; WC: Waist circumference; WHR: Waist-hip-ratio; DBP: Diastolic blood pressure; SBP: Systolic blood pressure; TG: Triglycerides; ALT: Alanine aminotransferase; AST: Aspartate aminotransferase; PLT: Platelet; CTRP-1: Complement c1q tumor necrosis factor-related protein 1; LDL: Low-density lipoprotein.

**Figure 3.**
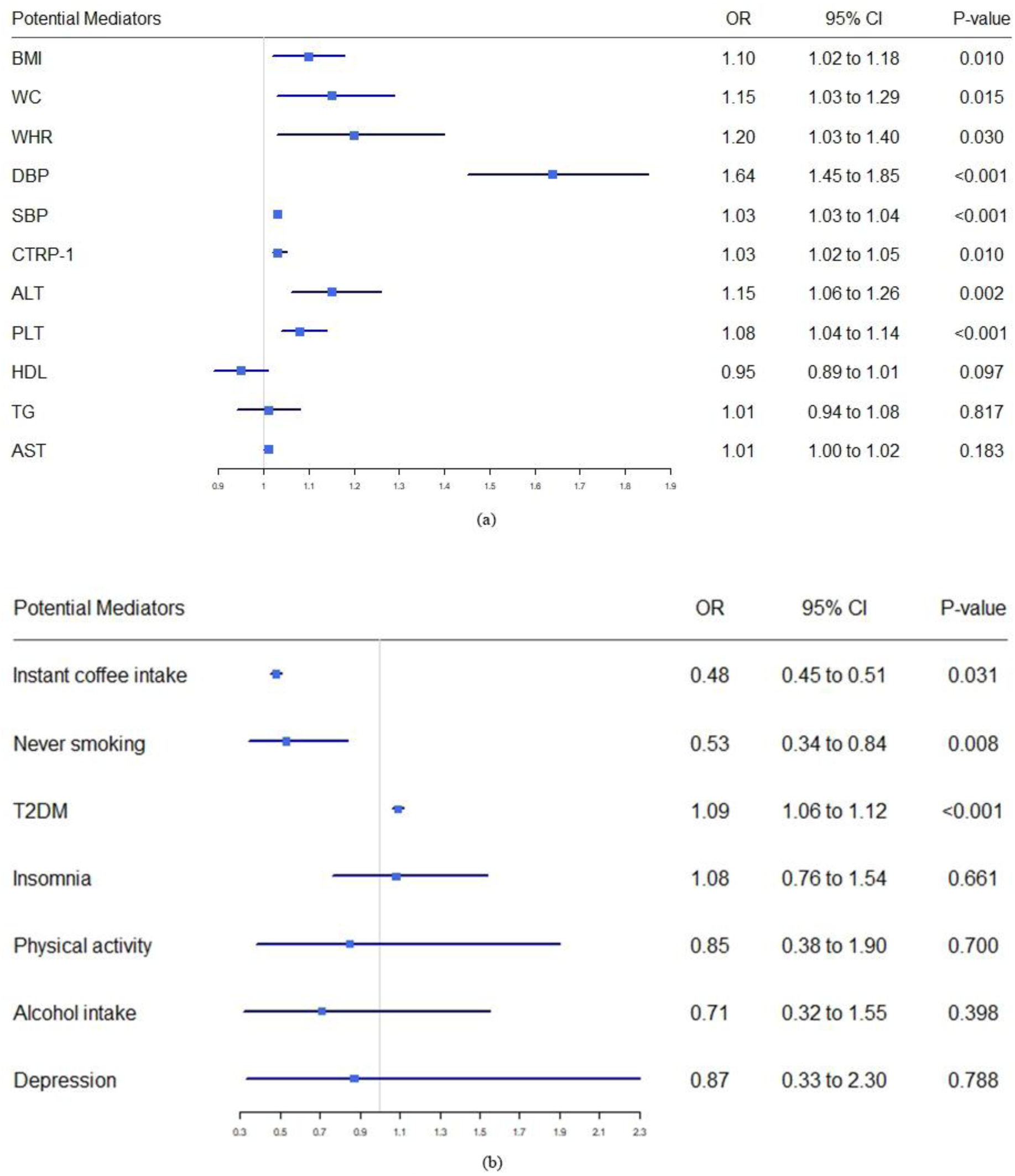
(a). Estimate of one-standard deviation higher of each potential mediator on stroke after adjusting for age completed full-time education. (b). Estimate of one-unit higher log odds of each potential mediator on stroke after adjusting for age completed full-time education. OR: Odds ratio; CI: Confidence interval; T2DM: Type-2 diabetes; HDL: High-density lipoprotein; BMI: Body mass index; WC: Waist circumference; WHR: Waist-hip-ratio; DBP: Diastolic blood pressure; SBP: Systolic blood pressure; TG: Triglycerides; ALT: Alanine aminotransferase; AST: Aspartate aminotransferase; PLT: Platelet; CTRP-1: Complement c1q tumor necrosis factor-related protein 1.

### Mediation Effects of Potential Mediators on Stroke

The proportion of the effect of education level on stroke mediated by BMI, WC, WHR, DBP, SBP, CTRP-1, ALT, PLT, never smoking status, and T2DM was 10%, 13%, 19%, 51%, 31%, 7%, 7%, 5%, 26%, 20%, respectively. Besides, the proportion mediated by instant coffee intake was -37%, which indicated that people with higher education level consumed less instant coffee, resulting in increased risk of stroke. The total mediation effect of continuous variables including BMI, WC, WHR, DBP, SBP, CTRP-1, ALT, and PLT was -0.238 (p < 0.05), which accounted for 79% of the effect of education level on stroke.

### Sensitivity Analyses

We performed multiple sensitivity analyses for two-sample MR results. Results of Weighted Median, MR-Egger, and MR-Egger intercept were shown in Supplementary Table 42. Results of MR-Egger regression indicated possible horizontal pleiotropies (p > 0.05 with nonzero intercept), however, p-values of these MR-Egger intercepts were all larger than 0.05, which showed evidence of pleiotropies, but their influence on our main analyses were not statistically significant. Moreover, results of another sensitivity analysis which aimed at detecting pleiotropy, MR-PRESSO, yielded similar results. Although results of MR-PRESSO showed some outliers, p-values of the MR-PRESSO distortion test were mostly larger than 0.05, thus indicated that differences of the causal estimates before and after removal of outliers were not statistically significant. Most results of weighted median were statistically significant (p < 0.05) which indicated that results of our study were less likely biased by invalid instruments. Results of other sensitivity analyses including single SNP analysis, leave-one-out analysis were consistent with our main analysis. Detail of all these sensitivity analyses were shown in Supplementary Table 42, 62-97.

## Discussion

The present large-scale MVMR study confirmed that higher level of education was causally associated with lower risk of stroke. More importantly, our results provided evidence that the effect of education level on stroke was mediated by 11 modifiable factors including lifestyle behaviors, cardiometabolic factors and other biomarkers. Taken together, all these mediators accounted for over 70% of the total effect. Thus, interventions on these factors might largely mitigate the risk of stroke.

Many previous observational studies have convinced that education level was associated with the risk of stroke (23–26). Additionally, higher genetically determined education level was also associated with reduced risk of stroke in MR studies (27–29). Although results from observational studies and MR studies were consistent, few studies have explored potential mediators of the protective effect of education level on stroke.

Our study was the first MR study exploring modifiable mediators of the causal effect of education level on stroke. Results of our study showed that intervention on lifestyle behaviors including instant coffee intake, physical activity, smoking, and cardiometabolic factors including BMI, WC, WHR, T2DM, DBP, SBP, ALT, PLT, CTRP-1 could reduce the risk of stroke. Thus, we suggested that people should try to attain higher level of education to reduce the risk of stroke, and for those who had difficulties in achieving higher level of education, intervention on those modifiable mediators might also have benefit in stroke risk reduction.

Results from previous observational studies exploring coffee intake and the risk of stroke were mixed (30, 31). Moreover, previous MR studies reported similar results that coffee intake was not causally associated with risk reduction of stroke(30, 32, 33). The negative results of these MR studies might be due to the combined effects of antioxidant effects(34, 35), inhibition aggregation of platelets(36, 37), and elevated plasma lipids(38, 39). We also noticed that none of these above MR studies categorized ground coffee and instant coffee, and interestingly, we found that people with higher education level tended to consume less instant coffee, and instant coffee intake was associated with less risk of stroke. A recent MR study also reported protective effect of instant coffee intake(40). Thus, results of our study suggested that more instant coffee intake for people with higher level of education might be helpful to reduce the risk of stroke. Our study found people with higher education level were more likely to be non-smoker, which contributed to the risk reduction of stroke. Previous observational and MR studies also reported protective effect of non-smoking status (41, 42). Based on results of previous studies and our study, non-smoking status was an important contributor to reduce the risk of stroke, and on the other hand, for people with low level of education, which was hard to improve, non-smoking status might be an essential modifiable mediator for reducing the risk of stroke.

Many observational and MR studies have convinced that cardiometabolic disorders including obesity, hypertension and T2DM were associated with increased risk of stroke(43–47). Results of our study also indicated that these cardiometabolic factors were modifiable mediators, and their mediation roles in the causality of education level on stroke were also essential.

Our study also indicated that several biomarkers might also be potential target, and intervention on these mediators could also reduce the risk of stroke. Kim et al. found that elevated level of ALT was associated with increased risk of intracerebral hemorrhage in Asia population(48). In contrast, Ford et al. reported opposite results in European population(49), Ruban et al. and Weikert et al. did not detect association between the level of ALT and the risk of stroke(50, 51). Results of our study indicated people with higher education level were associated with lower level of ALT, thus could reduce the risk of stroke. We speculated that lower level of ALT might be associated with poorer nutritional intake, thus was associated with reduced risk of stroke. But the inner mechanism of the relationship between the level of ALT and stroke remained controversial. Many previous studies have convinced that platelet adhesion, activation and aggregation were associated with hemostasis, which could finally lead to occlusion of cerebral vessels and stroke(52–54). Results of our study showed similar results, and reducing the level of PLT might be beneficial to risk reduction of stroke. Complement c1q tumor necrosis factor-related protein (CTRPs) family is a cluster of adipokines, and researchers have identified over 15 members (55), and CTRP-1 has been associated with cardiovascular risk (56). However, few researches have explored the relationship between CTRP-1 and the risk of stroke. Our study found CTRP-1 was positively associated with the risk of stroke, and people with higher education level were associated with lower level of CTRP-1, and thus with lower risk of stroke. Besides, we did not detect causal effect of insomnia, physical activity, alcohol intake, depression and level of HDL, TG, AST on stroke. Results of observational studies showed patients with stroke had higher rate of insomnia (57), and results from another meta-analysis showed insomnia could increase the risk of stroke(58). Observational studies were prone to be biased by confounding factors and thus were less conclusive in exploring causal effect, and the causality between insomnia and stroke was yet to be explored. A bi-directional MR study might be helpful to further explore such causality. Some researchers reported that physical activities could reduce the risk of stroke(59, 60), while other studies did not detect protection effect regarding more vigorous physical activities (61).

In the present study, genetic variants of physical activities were proxies for swimming or cycling, thus the level of physical activity might exceed the threshold for stroke prevention. Results from a previous MR study showed that heavy alcohol consumption could largely increase the risk of stroke, while the influence of light consumption was minimal (62). Limited by genetic instruments, we could not perform subgroup analyses according to the amount of alcohol consumption, and subgroup MR studies were expected in the future. Previous bi- directional MR study did not find causal correlations of depression with stroke (63), and depression was recognized as a common sequelae after stroke (64), thus more researches regarding the correlation between depression and stroke were needed in the future. Many researchers have found that HDL had protective effect on stroke, while LDL and TG could increase the risk of stroke (65–67), but results of some researches were conflicting (68). While the inner mechanisms remained largely unexplained, the correlation between lipid profiles and stroke might be multifactorial, which led to such paradox. Future exploration of various lipid metabolism pathways and other possible mechanisms might help further illustrate the above correlation.

We found that the combined mediation effect of mediators including instant coffee intake, physical activity, smoking, BMI, WC, WHR, T2DM, DBP, SBP, ALT, PLT CTRP-1 accounted for 79% of the total effect of education level on stroke. Restricted by many social, economic and other reasons, many people with low education attainment might have difficulties in improving education level after graduation, which led to excess risk of stroke. Thus, intervention on these factors might have benefit to risk reduction of stroke among these people with low education attainment.

## Strengths and Limitations

Several MR studies have identified that higher education level could reduce the risk of stroke(9, 10), but few had further explored mediators which could be used as intervention target. To the best of our knowledge, the present study is the first two-sample MVMR study to explore modifiable mediators of genetic correlation between education level and stroke. Influenced by confounders and measurement errors, interpretation of results from observational studies should be more cautious, and designing large-scale RCT was largely infeasible, MR studies which applied genetic variants as proxies, were more suitable. As genetic variants were determined at conception, results of MR studies were unlikely to be influenced by reverse causation, MVMR approach and sensitivity analyses could largely avoid bias caused by confounders and horizontal pleiotropy. Besides, we applied a two-sample method which could increase statistical power compared with one-sample MR method (69). In addition, genetic instruments used in our study survived LD and had large F-statistics which increased robustness of our results. Finally, we included as many possible mediators as possible to make our research more comprehensive.

The main source of bias for MR studies was horizontal pleiotropy, which affect approximately 48% MR studies (70). We applied multiple sensitivity analyses to detect horizontal pleiotropy. Most results showed null horizontal pleiotropy, and a few results indicated possible horizontal pleiotropy, but results of MR-Egger intercept and MR-PRESSO distortion test showed that bias caused such horizontal pleiotropy did not reach statistical significance threshold. The only exception was the estimate of effect of education level on DBP, MR-PRESSO distortion test and MR-Egger intercept reported conflicting results, which indicated the mediation effect of DBP might be somewhat influenced by horizontal pleiotropy. In addition, restricted by open-access GWAS datasets, we were unable to study the effect of education level on different stroke subtypes. Results of the present study reported null causal effect of insomnia, physical activity, alcohol intake, depression, and level of HDL, TG, AST on stroke after adjusting for education level. We found lack of convincing results from RCTs and lack of researches of mechanisms were main reason, and more RCTs, more subgroup MR analyses and more researches regarding inner mechanisms were expected in the future. Another limitation was the population included in our study was largely European which increased difficulties when generalizing our results to people with other ancestries.

## Conclusion

We applied a two-step, two-sample MVMR method and found education level had a protective effect on the risk of stroke. Further mediation analyses showed that through intervention on many modifiable mediators could also largely reduce the risk of stroke, which shed new light on stroke prevention.

## Data Availability

All data in the manuscript are available.

http://gwas-api.mrcieu.ac.uk/

## Acknowledgments

The authors acknowledge participants and researchers of multiple on-line GWAS databases for sharing summary-level data. Ken C and Ying Z contributed equally to the present study. Ken C and Ying Z designed this study. Ken C performed statistical analysis. Ken C and Ying Z wrote the first draft of the manuscript. Zheng W and Ying Yu reviewed the manuscript. All the authors contributed to the final version of the manuscript.

## Sources of Funding

NA

## Disclosures

None.

## Supplementary Materials

STROBE-MR Checklist Supplementary Tables

## Notes

### Competing Interest Statement

The authors have declared no competing interest.

### Clinical Trial

The present study is a Mendelian Randomization study, thus we did not have a clinical trial ID.

### Funding Statement

No external funding was received.

### Author Declarations

The present study was exempted from ethical approval because it used summary-level data.

